# WICID framework Version 1.0: Criteria and considerations to guide evidence-informed decision-making on non-pharmacological interventions targeting COVID-19

**DOI:** 10.1101/2020.07.03.20145755

**Authors:** Jan M Stratil, Maike Voss, Laura Arnold

## Abstract

1

**Introduction:** Decision-making on matters of public health and health policy requires the balancing of numerous, often conflicting factors. However, a broad societal discourse and a participatory decision-making process on the criteria underpinning the decision was often not feasible within the time constraints imposed on by the SARS-CoV-2 pandemic. While evidence-to-decision frameworks are not able or intended to replace stakeholder participation, they can serve as a tool to approach relevancy and comprehensiveness of the criteria considered.

**Objective:** To develop a decision-making framework adapted to the challenges of decision-making on national and sub-national level implementation of non-pharmacological interventions (NPIs) measures to contain the global SARS-CoV-2 pandemic.

**Methods:** We employed the “best-fit” framework synthesis technique and used the WHO-INTEGRATE framework Version 1.0 as a starting point. In a first phase adapted the framework through brainstorming exercises and application to exemplary case studies (e.g. school reopening). In a second phase we conducted a content analysis of comprehensive strategy documents intended to guide policymakers on the phasing out of applied lockdown measures in Germany. Based on factors and criteria identified in this process, we adapted previous framework versions into the WICID (*W*HO-*I*NTEGRATE *C*OV*ID-*19) framework Version 1.0.

**Results:** Twelve comprehensive strategy documents were included in the content analysis. The revised WICID framework consists of eleven criteria, supported by 48 aspects, the metacriterion *quality of evidence* and embraces a complexity and systems-perspective. The criteria cover implications for the health of individuals and populations due to and beyond COVID-19, infringement on liberties and fundamental human rights, acceptability and equity considerations, societal, environmental, and economic implications, as well as resource and feasibility considerations.

**Discussion:** In a third phase, the proposed framework will be expanded through a comprehensive document analysis focusing on key-stakeholder groups across the society. The WICID framework can be a tool to support comprehensive evidence-informed decision-making processes.

**Key-questions:** *What is already known?:* Ad-hoc Decision-making on matters of public health and health policy such as non-pharmaceutical interventions to contain the global SARS-CoV-2 pandemic, requires decision-makers to balance numerous and often conflicting criteria. Insufficient consideration of relevant factors reduces acceptance and can limit the effectiveness of the intervention.

*What are the new findings?:* Based on a content-analysis of comprehensive strategy documents, we newly developed WICID framework provides of 11+1 criteria informed by 47 aspects which are intended to support decision-makers in the balancing act of identifying and considering criteria of relevance.

*What do the new findings imply?:* The usage of the WICID evidence-to-decision framework can support decision-makers and expert committees in making more balanced decision, even if not all voices of relevant stakeholders could be included in the process due to time constraints imposed by the rapid progress of the pandemic.

## 3 Introduction

The response to the SARS-CoV-2 pandemic highlights the challenges of inherent evidence informed of public health and health policy decision-making^1-4^. These include: decision-making under time constraints, under uncertainty due to limited evidence, balancing numerous tradeoffs, and the challenge of ensuring fair decision-making processes under such circumstances.

Due to exponential growth in the number of infections, the issue of timing is crucial in a pandemic. A delay on the implementation of public health interventions (e.g. physical-distancing regulations) by days can have grave consequences^5^. Therefore, to receive timely evidence-informed guidance, many governmental institutions set up expert committees to support public health and health policy decision makers^6^. One challenge these expert groups faced is the qualitatively poor scientific evidence, with often questionable transferability and applicability to the context of decision-making and the lack of reliable evidence^7 8^. Not only due to the novelty of the pathogen, but also as assessing the effects of public health and health policy which often do not arise directly from the intervention, but from the system reaction to the introduction of the intervention within in it^9^ (e.g. school closures cause can lead to parents reducing working hours, which can lead to shortages in staff in the health care sector, limiting its ability to provide medical services and care).

However, even if strong scientific evidence was available, this in itself is insufficient to make sound recommendations, as evidence-informed decision-making is a deeply value-laden and often politized process^10-12^. Decision-makers must balance numerous and often conflicting normative and technical factors to come to a promising and acceptable decision^13-15^. This leads to the question: which criteria should be considered and how these should be weighed against each other?

The extent to which decisions are considered acceptable and legitimate depends on *how* the decisions were made (procedural considerations). Key considerations include e.g. transparency, inclusion of relevant stakeholders, and an appropriate composition of the panel^10 16-21^. Such approaches can increase the acceptability and perceived legitimacy of a decision^22-24^ even if – given varying and sometimes contradictory interests – no consensus regarding the *right* selection and weighting of criteria can be achieved^22^. According to the Accountability for Reasonableness (A4R) framework^20^, a key condition is ‘relevance’: the decision or recommendation must rest on evidence, reasons, and principles that all fair-minded parties can agree to be relevant and must meet the diverse needs of affected stakeholders^20^.

Involvement of representatives of all relevant stakeholder groups to identifying reasons and principles for a given decision-making process is considered ideal^10 16 25 26^. However, this ideal is often difficult if not impossible to meet under the time constraints imposed by the rapid progression of the SARS-CoV-2 pandemic. While not intended nor able to replace stakeholder participation, Evidence-to-Decision (EtD) frameworks are a way to support this balancing act^27 28^. EtD frameworks, which tend to comprise criteria and procedural guidance, are intended to ensure that all relevant factors are considered and the underlying rationale is made transparent^29^. When developed and applied well, these frameworks can help identify and integrate the criteria of relevance, even if not all voices of all stakeholders could be heard.

One of these frameworks is the WHO-INTEGRATE framework version 1.0^30^. It was developed in a research project commissioned by the World Health Organization (WHO) for complex public health and health system interventions^31^. Based on a conceptual and normative foundation^30^ primarily based on WHO norms and values derived and public health ethics frameworks^32-39^, it was developed based on a comprehensive literature review of real-world decision criteria^40^, an assessment of complexity features^30^ as well as qualitative research across four continents^41^. As with most EtDs, the WHO-INTEGRATE framework is generic framework. an requires adaption to the specific intervention and context.

## 4 Objective

The aim of this research project is to adapt the WHO-Integrate framework to support the development of recommendations and decision-making for lifting restrictions through non-pharmacological interventions (NPIs) to address the global SARS-CoV-2 pandemic. For this, the framework addresses the national and sub-national level and takes a plurality of viewpoints of affected stakeholders into account. The WICID framework (*W*HO-*I*NTEGRATE *C*OV*ID*-19 adaption) is intended to reflect decision-making challenges and opportunities on matters of public health in relation to COVID-19 by embracing a complex systems perspective. However, with the WICID framework, we aim to ensure that the tool is sufficiently generic to be applicable to a wide range of NPIs, contexts, and decision topics.

Although procedural criteria, norms, principles, and processual considerations are crucial for achieving fair processes, this research project focuses on the substantive decision-making criteria.

## 5 Methods

The development of the WICID framework was conducted in three phases, following an approach analogous to the “best fit” framework synthesis and using an adapted version of the WHO-INTEGRATE framework as a starting point^42^. An extended version of the methods used in this research project is provided as supplement 1.

In **phase I**, we adapted the WHO-INTEGRATE framework through brainstorming exercises and applying it to case studies, in order to develop an analytical, generic tool (a *priori* framework). This was done through discussion within the research team, (ii) assessment of real-world decision-making criteria derived from an comprehensive overview-of-reviews^40^, and (iii) conducting a brainstorming exercise guided by the application of the framework on two case studies (reopening high schools and reopening small businesses such as book shops). This preliminary *a priori* framework was then imported into the software MAXQDA20 (verbi, Berlin), with the criteria and sub-criteria of the a priori framework being translated into codes of the coding frame to be used in phase II.

**Phase II**, consisted of a content analysis of a purposive sample of comprehensive strategy papers on lifting the lockdown measures in Germany (e.g. reopening schools, increased testing measures), which were coded against the *a priori* framework developed in phase I following technique of "best fit" framework synthesis^42-44^. “Best fit” framework synthesis begins by creating a framework of *a priori* themes and coding data extracted from documents (in this case: the comprehensive strategy documents) against that thematic or conceptual framework. A new framework is created by performing a thematic analysis on any data that cannot be accommodated within the *a priori* framework^42^.

We assumed, that selecting comprehensive strategy documents by expert commissions or expert groups would provide a broad, multi-perspective set of recommendations (in contrast to e.g. scientific publications or statements by individual groups, which do not to reflect a broad range of relevant perspectives in conccluding). We defined these as documents (a) intended to provide a comprehensive strategy or strategic suggestions for phasing out the lockdown measures (rather than providing information or pointing out individual aspects), (b) not exclusively or primarily focused on mitigating the health related- consequences of the SARS-CoV-2 pandemic but also including other societal, economic, or health outcomes, (c) addressing various NPIs and their interplay, and (d) focusing on multiple considerations to be reflected in this process. Position papers of stakeholder groups reacting or positioning themselves to a document, measure, or event without providing comprehensive strategy guidance were excluded (n=8) but will be considered in upcoming **phase III**, where the current version of the WICID framework will be expanded (see 7.2). The eligibility criteria and the rationale for their selection are provided as a supplement in the expanded methods section.

The search was conducted through multiple pathways: two researchers (JMS, LA) independently conducted (1) grey literature searches in the search engine Google^TM^ and (2) on the websites of major newspaper outlets in Germany, (3) one researcher (JMS) searched the websites of the 16 German federal states, the national government and selected national government ministries, (4) we submitted freedom of information requests to the federal states’ governments, the national government, as well as selected national government ministries, and (5) contacted a sample of experts involved in public health decision-making or expert groups to provide us with strategy documents. As the documents are considered grey literature and mostly written in in German, we did not conduct a literature search in scientific data bases for these types of documents at this point of time.

The coding process of the included strategy documents was conducted by two authors (JMS, LA). Following the coding of one sample strategy documents to assess the need for adaption of the coding frame, one author (JMS) used the Software MAXQDA 20 (VERBI GmbH, Berlin) to code all identified strategy papers followed by a critical review by a second author (LA). The researcher applied the level 1 codes (referring to the *criteria* in the framework) and level 2 codes (referring to the *aspects* in the framework) of the coding frame to passages in the strategy document making references to criteria, considerations, or values covered within the codes. When the content of such a passage was perceived as not adequately covered by the coding frame, new level 2 or level 1 codes were created. Unclear passages were assigned the code TBD code for later review. The coding frame is provided as supplement 2.

After coding all selected strategy documents, two authors (JMS, LA) critically reflected on content saturation and dimensions of the framework insufficiently covered within the strategy documents and it was concluded that content saturation was reached.

Based on the coded passages, the researchers adapted the a priori framework by assessing the need to newly create, adapt, merge, separate, or reword *criteria* (reflecting level 1 codes) and aspects (reflecting level 2 codes).

In a final step, two researchers (JMS, LA) went through each of the coded passages to critically reflect on whether the criteria, considerations, or values contained within these passages was adequately covered within the newly adapted phase II framework.

## 6 Results

### 6.1 Development of preliminary adaption of the framework and a coding frame (phase I)

Following the process outlined in the methods section, we developed in phase I a preliminary, adapted *(a priori*) framework, which consisted of 8 *criteria* comprising 36 *aspects* and the metacriterion quality of evidence (an criterion to be applied across the other substantive criteria). This framework is provided as supplement 2. A preliminary version of this phase I framework as well as the exemplary case studies are part of a strategy document developed by two of the authors, among other experts, for the federal ministry of interior and the federal chancellery^45^.

### 6.2 Documents identified and included for analysis for phase II

We identified 12 comprehensive strategy papers in total. Four strategy papers, developed by expert groups for federal states, the national government, or ministries of the national government and two strategy documents developed by the corona expert commission of the federal state government of North Rhine-Westphalia, were publicly available^46^. Two strategy documents were developed by an informal expert group for the German Federal Ministry of the Interior, one of which was publicly available through a press report the other was provided to us through the German Network Public Health COVID-19 *(Kompetenznetz Public Health on COVID-19*)^45 47^.

We identified eight additional comprehensive strategy documents, which were not directly commissioned by governmental institutions. Four of which were published by the National Academy of Sciences Leopoldina^48-50^, one by a political party in the federal city-state of Hamburg^51^, one by a research institute commissioned by the political foundation Hans-Böckler-Stiftung^52^, one by the Boston Consulting Group^53^, and one developed by researchers from diverse institutions under the coordination of two researchers with affiliation at the University of Wuerzburg and the IFO institut^54^.

### 6.3 The adapted WICID framework

The adapted WICID framework consists of 11 substantive decision-making criteria, containing 48 decision-making aspects, and the meta-criterion quality of evidence, to be applicable across all criteria and aspects (outer circle, Figure 1; table 1 and 2). Depending on the measure, the criteria and aspects are intended to be applied on and reflected for different population groups (center-most circle, figure 1). Depending on the measure and type of decision-making process, the decision-makers are intended to deliberate on the criteria and aspects taking one or multiple different perspectives (inner circle, figure 1). Analogous to the WHO-INTEGRATE framework, it aims to accommodate different features of complexity: depending on impact the measure is assumed to have on the system it is implemented in, direct (those caused by the intervention) and indirect (those resulting from the system reactively changing due to the intervention) effects should be taken into account, as well as local, regional, national and even global implications. At the same time both, the immediate and the short, medium- and long-term implications should be considered (figure 2). It is intended to guide the systematic reflection of the intervention in its context.

**Figure 1:**
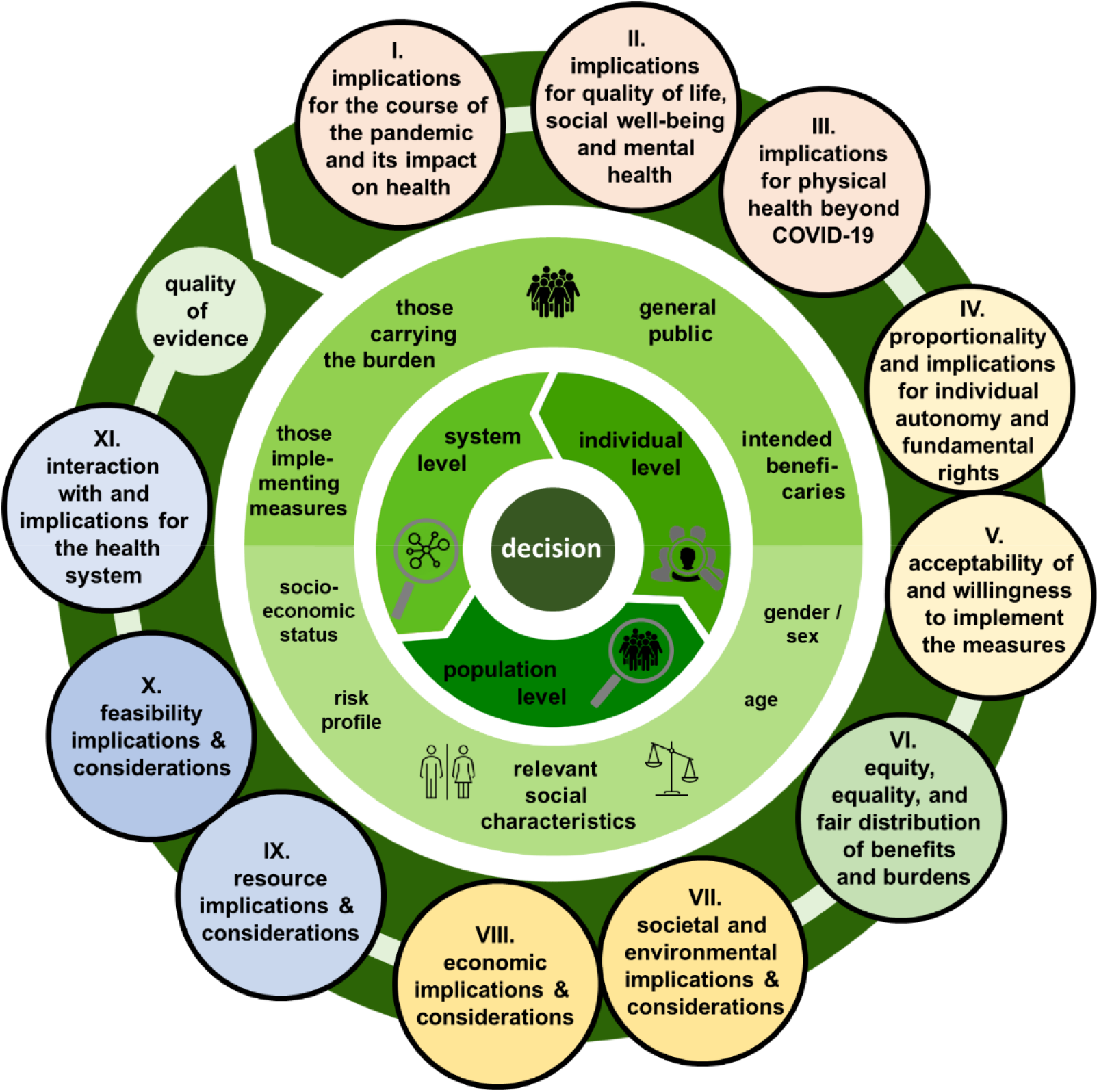
WICID Framework version 1.0 The color of the 11+1 criteria of the WICID refers so their grouping and relation to the criteria of the WHO-INTEGRATE framework they are derived from. The center-most circle describes population groups onto which the criteria and aspects should be applied to. The inner most circle describes the perspective the decision makers can take onto criteria and populations.

**Figure 2:**
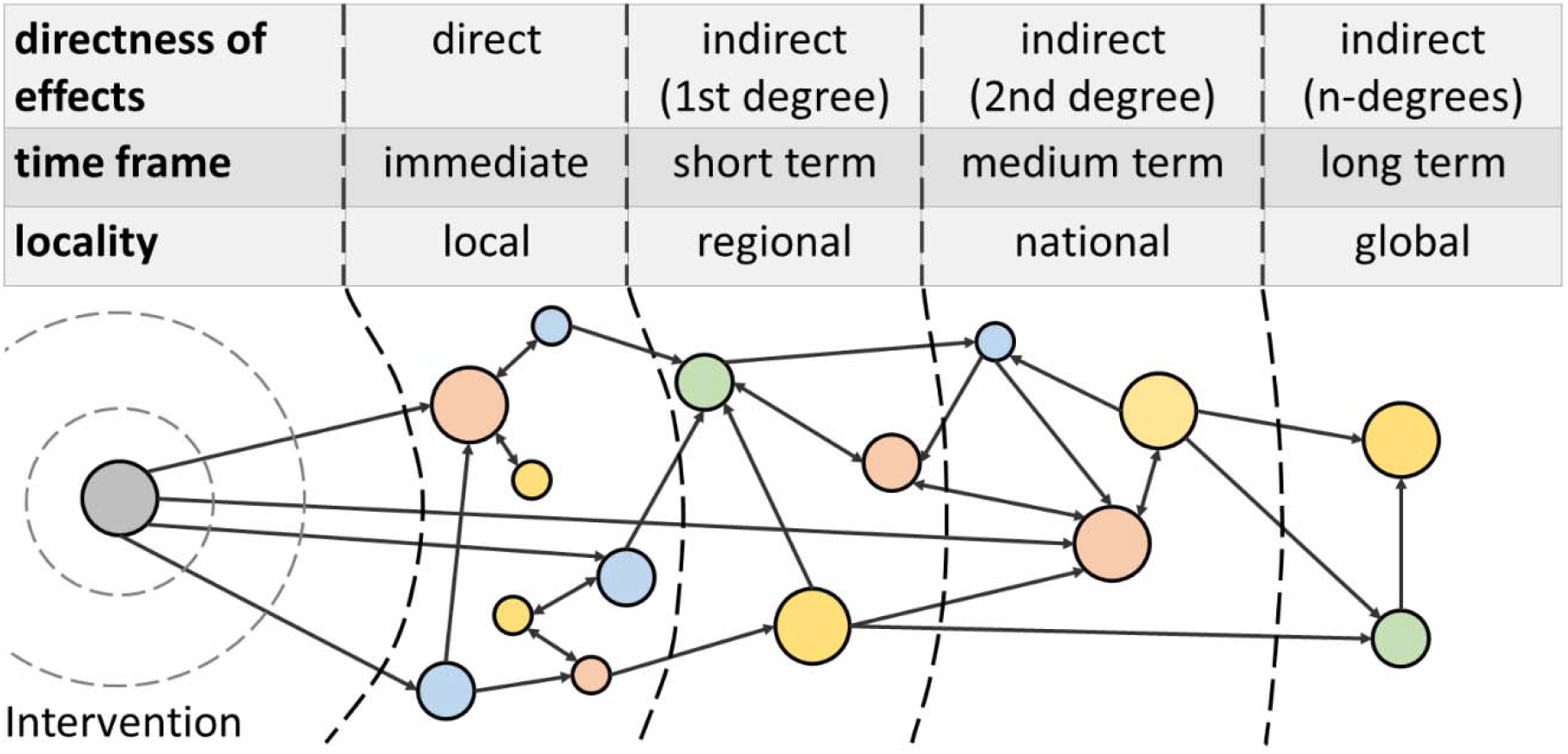
Complex system perspective on the implications caused by a measure being introduced into a system. The intervention (grey circle on the left) is introduced as an “event” to a system. It directly effects other components of the system (non-grey) circles on the right) which again interact with other components of the system, causing a chain-reaction of the system reacting and adapting to the event; leading to the societal, economic, and health-related consequences of the intervention.

#### 6.3.1 Criteria and aspects of the WICID framework

The eleven criteria in the WICID framework consist of: three criteria focused on the balance of health benefits and harms: (I-III, in figure 1: light red), two criteria focus on the accordance with human rights principles and socio-cultural acceptability (IV-V, beige), one criterion focuses on equity, equality, and non-discrimination (VI, green), two criteria focus on the societal implications (VII-VIII, light yellow), and three criteria focus on feasibility (IX-X, blue) and health systems considerations (XI, light blue). Table 1 describes the 11+1 criteria in detail; with table 2 containing the 48 associated aspects in their concise formulation. Supplement 3 contains a more comprehensive version of the framework, providing more details and examples to guide users of the framework. The supplementary 4 contains exemplary passages from the coded strategy documents for criteria and aspects.

**Table 1:** Criteria of the WICID framework and description of what the criteria are intended to cover

| **Criterion** | **Description of the criteria** |
| --- | --- |
| **I. implications for the course of the pandemic and its impact on health** | Covers implications of the measures for the course of the SARS-CoV-2 pandemic, including the number of new infections, the resulting health-related consequences (e.g. COVID-19 related mortality and morbidity), the implications for the capacity of the health care system to treat COVID-19 patients, as exceeding this capacity is associated with a pronounced increase in mortality. It furthermore covers the direct SARS-CoV-2 related health risk of individuals affected by the measures (probability of infection and consequences). |
| **II. implications for quality of life, social well-being, and mental health** | Covers the way the measures affect overall wellbeing and quality of life, which includes the degree to which the capability to shape everyday life according to one's own wishes and needs is affected (e.g. through restricting daily routines) or the experience of self-efficacy and of sense of coherence. It furthermore covers the social wellbeing of individuals (e.g. the experience of loneliness) and the social cohesion of communities (e.g. cohesion of families or non-family communities). The criterion furthermore captures the implications for the mental health of individuals and populations (e.g. depression, anxiety disorders), including risk-factors - such as the experience of stress or fear, ability to practice coping mechanisms, receive support, and other adverse mental health consequences (e.g. suicides). |
| **III. implications for the physical health, health behavior, health risks, and health care beyond COVID-19** | Focuses on behavioral, environmental, and interpersonal risk factors for health (including accidents and domestic violence) other than those directly related to COVID-19 (e.g. physical activity) and their consequences for health. It furthermore covers the implications of the measures for availability, accessibility, acceptability, and quality of medical and care services and institutions for conditions other than COVID-19 (e.g. willingness to seek emergency care in the case of myocardial infarctions). |
| **IV. Proportionality and accordance with individual autonomy and fundamental rights** | Covers whether the measure is in accordance with and how it – directly or indirectly – affects autonomy, self-determination, individual liberties, and fundamental individuals' rights (e.g. privacy and data protection implications of a contact tracing-app). It furthermore covers the intrusiveness of the intervention (with e.g. providing information being a measure with a low intrusiveness; and restricting choice one with a high intrusiveness<sup>37</sup>). The aspect of proportionality addresses whether intrusiveness and infringement of individual rights and liberties are proportionate to the expected benefit (or expected avoidance of harm). |
| **V. acceptance of and willingness to implement measures** | Focuses to the degree of acceptability of the measure and their consequences to the general population and different affected population groups. This includes the personal willingness to implement, adhere to, or enforce the measure (e.g. whether reopening schools without any protective measures is acceptable to teachers with a high-risk profile such as pre-existing health conditions). While acceptability is an end in itself, this criterion is strongly linked to feasibility and assumed effectiveness. |
| **VI. equity, equality and the fair distribution of benefits and burdens** | Covers the implications of the measure for vulnerable population groups, whether and how it affects stakeholder groups differently and thereby the risk for increasing or reducing inequalities (e.g. men benefiting less from a measure in comparison with women, exacerbating health inequities), considerations of equity (treating people differently according their need to allow them the same capability of achieving an outcome), considerations of equal treatment (e.g. not treating people differently without sufficient justification), and implications for the risk of individuals or population groups to be stigmatized or be discriminated against. |
| **VII. societal and environmental implications and considerations** | Covers the implications for civil society, social life, and culture from an individual-level and system-level perspective, and the implications for the functioning and cohesion of the society. It furthermore covers the implications of the measures for social determinants of health, including household income, social participation, and education. A further aspect |
|  | covered is the implications for the ecosystems resulting from the measures. |
| **VIII. economic implications and consideration** | Addresses the implications for the economy as a whole (population perspective/systems perspective) and of stakeholders in their role as economic actors and their activities (individual perspective. Including the implications of the measures for the work force (e.g. closing of schools forcing parents to stay at home to take care of their children). |
| **IX. resource requirements and resource implications** | Covers the requirements of the measures for different resources in the context of the availability of these resources as well as how the measure affect the availability and quality of these resources (e.g. how many face masks would be required to provide every teacher with a high-risk profile with one, are these masks available, and would this lead to a shortage of masks for e.g. health service providers). |
| **X. feasibility and implementation considerations** | Covers the practical, technical, and political feasibility of implementing the measure as well as the legal conformity. Other aspects address feasibility related characteristics of the measures, such as the flexibility in extension, adjustment, or withdrawal of the measures and the possibility of evaluating the implications of the measure and reacting adequately to new information (e.g. ability to test those affected by the measure and conduct contact-tracing if needed). |
| **XI. interaction with and implications for the health system** | Addresses how the intervention will interact (synergistically or adversely) with other measures to control and contain the SARS-CoV-2 pandemic and other not-directly COVID-19 related components of the health system (within a broad understanding of health system in accordance with the WHO<sup>55</sup>). For example, the combination of school reopening and lifting travel bans on other countries in close proximity timewise can have an interacting (negative) effect that could be different from each of these measures for lifting the lockdown by themselves. |
| **Quality of evidence** | This metacriterion is intended to be applied across all criteria and aspects, e.g. taking the quality of the evidence on health impacts alongside with its strength and direction into account. This metacriterion reflects the confidence that the available evidence is appropriate and adequate to support the decision or recommendation. Evidence is interpreted in a broad sense, beyond an understanding focusing on quantitative evidence of effectiveness derived from systematic reviews. Different forms of evidence can be used and be the most appropriate type of evidence to inform on the criteria (e.g. an appropriate form to assess the accordance with selected fundamental rights can be a legal assessment). Decision-making under uncertainty – as is often the case in a pandemic – often requires a decision based on stakeholder experience and judgement, when stronger evidence is unavailable. |

**Table 2:** Concise version of the criteria and aspects of the WICID framework

| criteria | aspect |
| --- | --- |
| I. implications for the course of the pandemic and its impact on health | implications for the **risk of infection** and **course of the pandemic** |
|  | implications of the measure for **COVID-19-related health consequences** |
|  | implications for the **capacity of the health care system** to treat (severe) COVID-19-cases |
| II. implications for quality of life, social well-being, and mental health | implications for the capability to **shape everyday life according to one's own wishes and needs** |
|  | implications for **individual wellbeing and quality** |
|  | implications of the measures for **social wellbeing** |
|  | implications for **social cohesion of communities** |
|  | implications for the **experience of fear and insecurity** |
|  | implications for **stress, stressors, the feeling of being overwhelmed and overloaded** |
|  | implications for **mental health and its consequences** |
| III. implications for the physical health, health behavior, health risks, and health care beyond COVID-19 | implications for **health-related behavior** |
|  | implications for the **exposure to environments that affect health** |
|  | implications for the **risk of suffering accidents or being affected by violence** |
|  | implications for **medical emergency and standard care beyond COVID-19** |
|  | implications for **care and other health and social support services** |
|  | **Other health related implications** beyond COVID-19 |
| IV. Proportionality and accordance with individual autonomy and fundamental rights | implications for and accordance with **individual liberties and fundamental rights** including **autonomy** and **individual self-determination** |
|  | extent of **intrusiveness** and **proportionality** regarding the expected benefit |
| V. acceptance of and willingness to implement measures | **Socio-cultural acceptability** of the measures |
|  | implications for **willingness to adhere to implement the measures** |
| VI. equity, equality and the fair distribution of benefits and burdens | implications for **health, social and economic inequalities** |
|  | **implications for affected social groups**, in particular **vulnerable and marginalized populations** |
|  | degree of **fair distribution of benefits and burdens** between population groups |
|  | **accordance of the measures with the principle of equal treatment** |
|  | implications for the risk of **stigmatization and discrimination** |
| VII. societal and environmental implications and considerations | **implications for actors and institutions of civil society, social life, and culture** |
|  | **implications for the civil society, social life, and culture** |
|  | implications for **social cohesion, solidarity, and the risk of social and political division** |
|  | implications for **the economic situation of individuals and capability of social participation** |
|  | implications for **education and psychosocial development** |
|  | implications for other **social determinants of health** |
|  | implications for **ecosystem, planetary boundaries, and environmental sustainability** |
| VIII. economic implications and consideration | **consequences** of the measures for **individual economic actors** and their economic activities |
|  | **implications for the economy** including resilience and ability to recover |
|  | **implications for innovation and economic development opportunities** |
|  | **implications of the measures on the retention and release of labor forces** |
| **IX. resource requirements and resource implications** | **financial costs, available financial resources, and budgetary implications for different actors** |
|  | **requirements of, availability of, and implications for human resources** |
|  | **requirements of, availability of, and implications for protective equipment, hygiene products and medical devices** |
|  | **requirements of, availability of, and implications for infrastructure to implement, enforce and monitor measures** |
|  | **requirements of, availability of, and implications for further resources and infrastructures** |
| **X. feasibility and implementation considerations** | **political feasibility and legal conformity** |
|  | **practical and technical feasibility** |
|  | **flexibility in implementation, extension, adjustment and withdrawal of measures** |
|  | **possibility of evaluation and adequate reaction to new information** |
| **XI. interaction with and implications for the health system** | **interaction with other measures to control and contain the SARS-CoV-2 pandemic** |
|  | **interaction with components of the health care system** |
|  | **implications for the capability and willingness to develop alternative, local solutions** |
| **Metacriterion: quality of evidence** |  |

#### 6.3.2 Considering criteria for different populations

Depending on the measure and decision-making context, criteria and aspects within the framework should be considered for the population as a whole, as well as for different population groups to assume relevant implications adequately for these groups. For example, an intervention such as regulation forcing people not to leave their houses can have adverse effects which disproportionate affect people affected by insecure housing circumstances or school closures disproportionately placing a burden on working mothers. The stakeholder groups to be considered will depend on the type of measure (e.g. closing schools vs. closing nursing homes to the public). Building on the WHO-INTEGRATE framework and the strategy documents, we suggest to consider the implications for (figure 2, center-most circle): (a) the general population, (b) those intended to benefit from the intervention (e.g. young school children in the case of school reopening), (c) those intended to implement the measures (e.g. teachers), and populations with a high risk profile (senior citizens with preexisting conditions) and (f) other affected stakeholder groups (e.g. employers). Within these population groups, further disaggregation based on relevant social characteristics with an emphasis on vulnerable and marginalized populations should be conducted (e.g. school children with a family with a low socio-economic status). These relevant characteristics are likely to vary depending on context, although socioeconomic status, age, and gender are likely to be important across context. The PROGRESS Plus framework can provide guidance on the identification of relevant characteristics^56^.

#### 6.3.3 Employing multiply perspectives: The population, the individual and the health system

Depending on the measure and decision-making process, decision-makers need to reflect on the different criteria from different perspectives to inform their deliberations (inner ring in figure 1). For example, the health implications of a measure on the SARS-CoV-2 related health risk (e.g. reopening of schools) can be approached from a population perspective (looking at the implications of the intervention for the population as a whole, which can take place on a local or community level, federal state level and/or national level; e.g. incidence rate of infections and associated mortality rates), a systems perspective (reflecting the intervention from the perspective of the health system, as well on a local, regional, and national level, e.g. taking the implications for the capacity of the health system and the availability of resources into account), or the individual perspective (taking the perspective of an individual affected by the population; e.g. the risk for individual teachers working in these schools).

#### 6.3.4 Taking a complex systems perspective

The implications of the NPIs can reach far. Therefore, the WICID framework embraces a complexity perspective^2^: the measures are regarded as “events in a system”^57^, with the (intended and unintended) effects resulting from the interaction of the measures’ components with each other and components of the larger system. Within this perspective, disentangling the effects *caused by* the measure itself from the interplay of context and measure can be challenging if not impossible to do^9^; posing challenges for transferability and generalizability of evidence.

Analogous to the ripple effects caused by dropping an object in a pool of water, the effects initiated by the introduction of the measure to the system can lead to a chain reaction that can be followed outwards incrementally (figure 2). The more profound the impact of the measure to the system, the further way the effects of the measure throughout the system can be observed. For example, the shutdown of a few companies in a region for a short time can lead to locally felt adverse economic consequences. However, a marginally more impactful event of closing the same companies for a marginally longer time and thereby exceeding an economic threshold can lead to the insolvency of these companies, causing – depending on the companies – a disruption of globalized production chains with economic consequences that can have regional, national, and even global effects. Therefore, depending on how profound the impact of the measure is assumed, decision makers need to consider whether the measure is likely to lead not only to immediate and local, but also regional, national, or global consequences over the short, medium, and long term. When reflecting on the measures, a focus should not only be on the direct effects along the intended causal pathway, but should also anticipate implications caused across several degrees of indirectness. These different dimensions are not necessarily related: for example, direct and indirect health-related, societal, and economic implication occurring from immediate to long term could arise exclusively on a local level (figure 2).

## 7 Discussion

### 7.1 Summary of findings

We adapted the WHO-INTEGRATE framework to decision-making processes on non-pharmaceutical interventions intended to suppress or mitigate the effects of the SARS-CoV 2 pandemic. We used brain storming exercises and content analysis of comprehensive strategy papers on the phasing out the implemented lockdown measures in Germany. The resulting WICID framework version 1.0 consists of 11+1 substantive decision-making criteria, containing 48 decision-making aspects. Depending on the needs of the decision-making processes, these are intended to be applied by policymakers for and with different affected stakeholder groups using a multi-perspective approach. In line with the underlying complexity perspective, rather than only focusing on direct, immediate, and local effects of the measure, the ripple effects caused by the introduction of the measure to a given system or policy field should be followed to adequately consider the implications a measure might have.

### 7.2 Advancing the WICID framework version 1.0 in phase III

The current version of the WICID framework version 1.0 will be expanded in a third phase. The limited diversity of expert groups established to inform policymakers on the handling of the SARS-CoV-2 pandemic has faced some criticism^6^. Our approach of adapting the WICID framework based on strategy documents therefore comes with the risk that relevant criteria were overlooked due to the limited selection of expert groups and the stakeholder groups (not) represented within them. The third phase of the research project aims to address this issue by including the perspectives of various stakeholder groups across the society and expanding the WICID framework version 1.0 with considerations not adequately covered previously.

In phase III we will conduct a content analysis of key-documents representing the opinions and perspectives of stakeholder representatives across the society (i.a. of affected populations, non-governmental organizations, private sector) and using the results to validate and – where needed – expand the framework version 1.0. Using a sample of NPIs with broad societal implication as a starting point (closure/reopening of schools, closure/reopening of businesses, and “shelter-in-place” regulations), we will include opinion pieces, position papers, or press statements aimed at informing political decision-making on these measures. A first set of stakeholder group clusters (e.g. social and welfare organizations) will be selected based on an initial brainstorming phase and stakeholder mapping^58^ and expanded in an iterative snowballing process. While it will be not feasible to cover all relevant organizations within a given cluster, we will analyze of a heterogeneous sample which will be expanded in an iterative process based on the assessment of saturation. While this approach is not able to capture all voices of affected stakeholders, it allows for a broad and representation of societal values in decision-making across the.

### 7.3 Short guidance on how to apply the WICID Framework

While some evidence-to-decision (EtD) frameworks are tailored to specific decision-making processes (e.g. on vaccination policies^59^) and provide a fixed set of “ready-to-use” decision-making criteria^26^, others are more generic and require some form of adaption. The WICID framework is intended to be adequately generic to be applicable across a broad range of NPIs and decision-making contexts. While the 11+1 criteria can be used as a “ready-to-use” evidence-to-decision framework, we believe the framework to be most useful as a guide to systematically reflect on NPIs as well as their interdependencies and adapting them based on the specific needs of the decision-making process, using the WICID framework as guidance.

First, (I) a comprehensive logic model^60 61^ or systems map of the measure and the context is intended to be implemented in should be created, in order to describe possible implications. Next, (II) the WICID framework should be used to expand on dimensions not adequately covered (e.g. by exploring the causal pathways from different perspectives, assessing the implications for different affected population groups, or using the criteria and aspects to assess its comprehensiveness). Informed by the logic model, (III) an identification of relevant stakeholders should be conducted, ideally in the form of a comprehensive or focused stakeholder mapping^58^. Next, (IVa) those involved in the decision-making process need to define criteria which are assumed to be of relevance for deliberating on the measure. This can be done e.g. by selecting individual aspects from within each criterion and adapting them to the context at hand. Using the example of school reopening, this could include the risk of outbreaks, health implications for teachers, for students, and for family members, the implications for the wellbeing of these groups, educational implications etc. Given the complexity of the decisions at hand, it is likely not all factors of relevance can be covered in depth. On the one hand, this reflects the reality of the decision-making process; on the other hand, the rationale for the selection should always be provided. (IVb) The assumed importance of the criteria should be rated (e.g. on a 1-5 scale from “less important” to “critical”). (V) Efforts should be made to receive feedback on the expanded logic model and the selected criteria from key stakeholder groups identified in the mapping. Repeated rounds of step I-IV are likely to produce the best results. Next (VI), efforts should be made to acquire appropriate sources of evidence to inform on the selected criteria (e.g. by commissioning research or inviting experts’ judgments). (VII) The retrieved evidence for each criterion should be summarized and presented alongside the assessment the quality of the evidence and transferability to the context at hand. The group of decision-makers are now asked (VIII) to engage in the deliberation to balance the criteria against each other, taking their weight, direction, quality, and transferability of the evidence into account. After all, the final judgment and the underlying rationale should be made transparent and public.

### 7.4 Relation to public health ethics framework

Various public health ethics frameworks providing guidance on principles and values to consider in public health and health policy decision-making have been published^39 62-64^, some of which are more generall^33 34^ ^37 65 66^, while others focused on public health emergencies and pandemics^67-71^. Building on these foundations, institutions such as the German ethics council^72^ or the German Network Public Health COVID-19^73 74^ have outlined relevant values and principles for decision-making in the current public health crisis. These include the duty to provide care, health, non-discrimination, security, equity, individual liberty, privacy, proportionality, protection of the public from harm, reciprocity, and solidarity, among others^73^.

The WICID framework was developed to be in line with these documents; primarily due to the underlying WHO-INTEGRATE framework being developed with a foundation in WHO norms and values and key public health ethics frameworks^30^. The WICID framework aims to translate these principles and values into criteria applicable for real-world decision making processes in the pandemic (e.g. by translating the general moral considerations of producing benefits and avoiding, preventing, and removing harms^33^ into – among others – the criteria I,II,III,VII, and VIII on the different positive and negatives social, economic, or health-related effects a NPI might have for individuals and populations). Furthermore, the framework aims to place criteria derived from these values and principles alongside other factors of relevance for real-world decision-making often not covered in depth in public health ethics frameworks, such as considerations of feasibility or the wider implications for the (health) system. While some values and principles have a direct representation in the framework criteria (e.g. individual liberty, privacy, or proportionality) others are introduced on the level of the perspectives or the populations the criteria should be applied to (e.g. reciprocity being reflected in the consideration of *those intended to implement the interventions)*.

### 7.5 Need for fair and transparent processes

The WICID framework is in itself insufficient to achieve fair decision-making processes with results considered acceptable and legitimate^20 21^. It is important that not only the final decision, but also the underlying rationale, including the criteria and evidence, is made transparent. Other important values and principles underlying fair decision-making processes include accountability, inclusiveness, openness and transparency, reasonableness, and responsiveness^73 74^. While a comprehensive approach of stakeholder engagement is likely not possible within a pandemic, efforts to including the voices of affected stakeholders, for example through rapid reaction statements from stakeholder representation organizations, can not only improve acceptability and legitimacy but also lead to better outcomes. Especially, the needs of affected marginalized and vulnerable population groups without strong political capital (e.g. people affected by homelessness) are at risk of being overlooked. Furthermore, special attention needs to be placed to the composition of the stakeholder groups^6^. Documenting the process and the decisions made and providing – to some extend – access to these documents to the public can increase social acceptance for public health measures as well as lifting them. As the main focus within this research project lay on substantive criteria of WICID the framework and does *not* comprise procedural criteria, other procedural frameworks e.g. from the field of public health ethics can serve as guidance.

### 7.6 Strengths and Limitations

The WICID framework was developed by building on the WHO-INTEGRATE framework^30^ which was developed as a principles-based approach to ensure a solid, comprehensive normative foundation. It is also based on previous research such as the result of an overview of systematic reviews on public health and health system decision-making criteria^40^, and expanding this foundational framework through a broad set of comprehensive strategy documents informing decision-making processes in Germany. Application in other country contexts therefore need to be tested and the framework, if necessary, updated and revised.

The WICID framework was developed using strategy documents intended to inform the German government. While we believe the resulting WICID framework can prove useful and applicable to other regions within and outside Europe, the need for adapting to the respective decision-making contexts is necessary. Likely, the basis of the WHO-INTEGRATE framework, which was developed not only for the global level at the WHO, but also to be applicable on national and sub-national levels throughout the world, can cover factors not adequately captured in the German strategy documents.

The framework in its current version 1.0 was developed based primarily on comprehensive strategy documents developed by expert groups. The composition and the intention of the expert groups is likely to have influenced the criteria, consideration, values, and principles covered within them. While we believe possible shortcomings and blind spots are in part compensated using the WHO-INTEGRATE framework as a basis, there is a risk of relevant factors being missed. We aim to address this issue in phase III of this research project.

Despite multiple approaches to identify the comprehensive strategy documents, we acknowledge the possibility of having missed on individual statements. Furthermore, likely other relevant strategy documents exist, but were not disclosed by the governments or leaked through to the public and therefore are not captured in our analysis. An update of the searches will be conducted as part of phase III.

Another limitation is the distinction between comprehensive strategy documents and position papers by affected stakeholder groups, which was not always a clear cut. However, we believe this only to be a minor limitation, as all borderline documents were retained and will be included in the phase III of the project.

## 8 Conclusion

The WICID framework represents a comprehensive COVID-19 focused EtD framework intended to guide policy and public-health decision-makers on making decisions on NPIs. It is rooted in WHO norms and values, criteria and considerations used to inform decision-making, and a complex systems perspective. While adapted to COVID-19 related challenges, it is intended to be generic in a way to be applicable across a broad range of decisions-making processes, contexts, and on a diverse set of measures. It can be a useful tool for those involved in the difficult task of making decisions on NPIs by systematizing the decision-making process, making the underlying rationale more transparent and contributing to the relevance of the decision criteria.

## Data Availability

The included comprehensive strategy documents are - for the most part - publicly available. Those that are not, can be provided by the authors uppon request

## Acknowledgements

We would like to thank Prof. Eva Rehfuess for her comments and support in the development of the phase I framework. We furthermore would like to thank Prof. Rob Baltussen, Prof. Georg Marckmann, Karin Geffert, Dr. Lisa Pfadenhauer, Dr. Kerstin Sell, and Prof. Hajo Zeeb for taking the time to critically reviewing and commenting on the manuscript and framework, despite the time constraints the current pandemic imposes on them.

## Author’s contribution

JMS and LA conceived the study. JMS, EAR, MV and LA engaged in the activities of phase I to develop the a priori framework. JMS and LA conducted the literature search for strategy documents, with support from MV. JMS and LA conducted the coding and analysis of the strategy documents and revised the a priori framework. JMS drafted the manuscript with support from LA and MV.

## Patient consent

Not required.

## Ethics approval

The research will be undertaken in accordance with the declaration of Helsinki in their respective current versions. As it is a document analysis, no review by an ethics committee was deemed necessary.

## 10 Annex

### Supplements 1 – Expanded methods description

The development of the WICID framework was conducted in three phases, following an approach analogous to the “best fit” framework synthesis and using an adapted version of the WHO-INTEGRATE framework as a starting point^42^. In phase I, we adapted the framework through brainstorming exercises and applying it to case studies, in order to develop an analytical, generic tool *(a priori* framework). Phase II, consisted of a content analysis of a purposive sample of comprehensive strategy papers on lifting the lockdown measures in Germany (e.g. reopening schools, increased testing measures), which were coded against the *a priori* framework and based on which the WICID framework version 1.0 was created. In a next step (phase III), which is yet to be completed, we will advance the WICID framework version 1.0, by integrating the perspectives of a diverse set of affected stakeholders across society. Phase III allows for an assessment of the comprehensiveness of the framework criteria and the integration of factors insufficiently covered in the expert-based strategy documents.

#### 10.1 Phase 1: Development of a preliminary adaption of the framework and of a coding frame

In phase I, we (i) discussed the WHO-INTEGRATE framework within the research team, (ii) assessed real-world decision-making criteria derived from an comprehensive overview-of-reviews^40^, and (iii) conducted a brainstorming exercise guided by the application of the WHO INTEGRATE Framework version 1.0 on two case studies (reopening high schools and reopening small businesses such as book shops).

This preliminary *a priori* framework was then imported into the software MAXQDA20 (verbi, Berlin), with the criteria and sub-criteria of the a priori framework being translated into codes of the coding frame to be used in phase II. In this process, we added two additional codes: (1) “evidence” – to cover considerations regarding information and evidence considered necessary of lacking in the decision-making process, and (2) the code “TBD” *(to be discussed)*, which was meant mark unclear passages for later review and discussion by the research team.

#### 10.2 Phase 2: Coding of strategy documents, best-fit framework synthesis, and development of the WICID framework

We used the technique of "best fit" framework synthesis^42-44^, which offers a method to build on an existing framework that is considered as relevant for the given circumstances, but potentially different in relevant areas (such as the WHO-INTEGRATE framework for decision-making on COVID-19 related decision-making). “Best fit” framework synthesis begins by creating a framework of *a priori* themes and coding data extracted from documents (in this case: the comprehensive strategy documents) against that thematic or conceptual framework (see 8.2.1). A new framework is created by performing a thematic analysis on any data that cannot be accommodated within the *a priori* framework. We used the adapted version of the WHO-INTEGRATE framework created in phase I, and translated this into the coding frame we used coded the strategy documents against (provided in as supplement 2)^42^.

##### 10.2.1 The strategy documents and the rationale behind their selection

We assumed, that selecting comprehensive strategy documents by expert commissions or expert groups would provide a broad, multi-perspective set of recommendations (in contrast to e.g. scientific publications or statements by individual groups, which do not claim nor intend to reflect multiple relevant perspectives in concluding).

We defined these as documents (a) intended to provide a comprehensive strategy or strategic suggestions for phasing out the lockdown measures (rather than providing information or pointing out individual aspects), (b) not exclusively or primarily focused on mitigating the health related-consequences of the SARS-CoV-2 pandemic but also including other societal, economic, or health outcomes, (c) addressing various NPIs and their interplay (e.g. not exclusively focusing on testing), and (d) focusing on multiple considerations to be reflected in this process (i.e. not exclusively focused on health impact). Position papers of stakeholder groups reacting or positioning themselves to a document, measure, or event without providing comprehensive strategy guidance were excluded (n=8) but will be considered in phase III.

While the main discourse on implementing lockdown measures in Germany was focused on suppressing the spread of the outbreak and on averting a collapse of the health care system, the debate on the controlled phasing out of the implemented lockdown measures was more nuanced: focusing on the challenge to balance the implications of the measures e.g. on health, society, or the economy. Therefore, we concentrated on strategy documents that focused on the latter.

To include strategy documents with impact on real world decision-making, we selected papers developed by expert groups or task forces commissioned by German policymakers on two levels (federal and national governments as well as ministries on these levels). We expanded the sample to comprehensive strategy documents on the exit from the lockdown measures developed by non-government affiliated expert groups intended to inform political decision-making, but not directly commissioned by governmental bodies.

In order to ensure that the strategy documents addressed a broad range of societal and economic implications beyond the health sector, we focused on strategy documents which – directly or indirectly – addressed one of the following NPIs: (a) closure and reopening of schools, (b) closure and reopening of businesses, and (c) “shelter-in-place” regulations. We excluded strategy papers that were not or only marginally concerned with these three NPIs. Strategy papers primarily concerned with hygiene measures or the testing capacity were excluded as well.

Table S1 in the annex displays the inclusion and exclusion criteria for phase II. All identified borderline cases were retained to be analyzed in phase III.

**Table S1:** Inclusion and exclusion criteria for the search of eligible documents for phase II

| Inclusion criteria | Exclusion criteria |
| --- | --- |
| • The document addresses measures related to the handling of the SARS-CoV-2 pandemic in Germany<br>• The document is intended to inform real-world policy and public health decision-makers in Germany<br>• The document is intended to provide a comprehensive strategy or strategic suggestions for phasing out the lockdown measures<br>• The document addresses societal, economic, or health considerations beyond health considerations associated with SARS-CoV-2 (e.g. number of infections) | • The document is not primarily targeted to inform policy or public health decision makers<br>• The document does not provide a comprehensive strategy to phase out the lockdown, but is rather a reaction or positioning paper to an event, decision, or measure or provides general statements on aspects to consider (e.g. statement not to forget migrant workers)<br>• The document is only or primarily concerned with direct health consequences of COVID-19 and/or the mitigation of these effects (e.g. the right number of tests to be performed), |
| • The document focuses addresses multiple considerations to be considered in this process (e.g. not exclusively focused on health impact considerations).<br>• The document addresses considerations for multiple NPI or considerations of relevance for multiple NPIs on a more abstract level<br>• Position papers of stakeholder groups reacting or positioning themselves to a document, measure, or event without providing comprehensive strategy guidance are considered in phase III.<br>• To document was developed by an expert group or task forces implemented by or commissioned to write the document by the federal or national governments as well as ministries of the German federal Government OR by non-government affiliated expert groups intended to guide public health and health policy decision making on the federal or national level.<br>• The document addresses – directly or indirectly – addressed one of the following NPIs: (a) closure and reopening of schools, (b) closure and reopening of businesses, and (c) “shelter-in-place” regulations. | without discussing broader considerations (e.g. regarding societal implications, economic implications, feasibility and resource considerations)<br>• The document is exclusively focused on the effectiveness of a measure (e.g. impact of school closures on COVID-19 transmission), without taking broader considerations (e.g. societal, economic, feasibility, or ethical considerations) into account.<br>• The document is providing general information on COVID-19, the pandemic, or NPIs; not intended to guide decision-making (e.g. what is allowed in a federal state)<br>• The document is a scientific study, guidance documents and guidelines to inform patients or health care providers on treatment and therapeutic approaches of COVID-19 (e.g. guidelines on the safety of ibuprofen)<br>• The document is focused on health care system and medical care planning without relation to the selected NPIs (e.g. on triage-procedures, necessity of intubation therapy, relation of intensive vs. palliative treatment, number of ICU-beds necessary) |

#### 10.2.2 Identification and selection of strategy papers

The search was conducted through multiple approaches channels:

- Two researchers (JMS, LA) independently searched in the search engine Google™ with various versions of keyword combinations of the terms and synonyms of “strategy” or “expert commission” and “COVID-19” in German.
- Two researchers (JMS, LA) independently searched the websites of major newspaper outlets in Germany (including: Die Zeit, Frankfurter Allgemeine Zeitung, Die Welt, Deutschlandfunk, Der Spiegel) outlets using the website’s search engine with similar keywords
- One researcher (JMS) searched the websites of the 16 German federal states, the national government and selected national government ministries, focusing on the section of press releases.
- We contacted a sample of experts involved in public health decision-making or expert groups to provide us with strategy documents; either directly or through the platform of the interdisciplinary *Kompetenznetz Public Health COVID-19 (Competence Network Public Health COVID-19, www.public-health-covid19.de*).
- We posed freedom of information requests to the federal states’ governments, the national government, as well as selected national government ministries to provide us with strategy documents developed by expert groups, if available and publicly accessible.

As the documents are considered grey literature and mostly written in in German, we did not conduct a literature search in scientific data bases for these types of documents at this point of time.

#### 10.2.3 Coding of documents against the *a priori* framework

The coding process was conducted by two authors (JMS, LA; personal characteristics in line with the COREQ-checklist^75^ are reported in table S2). First, one sample strategy document was coded by the two authors to assess the need to adapt the preliminary coding frame and to develop a coding guidance document, outlining when a specific code should be used and which text passages to code. The coding frame (translated from German) is provided as a supplement (supplement 2). Based on this adapted frame, one author (JMS) used the Software MAXQDA 20 (VERBI GmbH, Berlin) to code all identified strategy papers. Afterwards, all coded documents were critically reviewed by a second author (LA) who highlighted conflicts (passages where this reviewer perceived a code was missing, suggested not to code, or to code differently). Conflicts were solved through discussion between the authors.

In the subsequent process, the researchers applied the level 1 codes (referring to the *criteria* in the framework) and level 2 codes (referring to the *aspects* in the framework) of the coding frame to passages in the strategy document making references to criteria, considerations, or values covered within the codes. Level 2 codes *(aspects)* are meant to describe considerations (e.g. factors, values, norms) contained within the level 1 codes *(criteria)* and support the user in the understanding, interpretation, and application of the criteria. When the content of such a passage was perceived as not adequately covered by the coding frame, new level 2 or level 1 codes were created. When the researchers identified passages containing references to criteria, considerations, or values of relevance which were assumed to be covered by a specific level 1 or level 2 code, but which seemed to expand on this or provide details or nuances (e.g. a passage on the implications of an measure risking the insolvency of small enterprises within the code *economic implications)*, the researchers took note of these passages for later review. Unclear passages were assigned the code TBD code for later review.

After coding all selected strategy documents, both authors critically reflected on content saturation and dimensions of the framework insufficiently covered within the strategy documents. As most criteria of the preliminary framework were adequately covered and due to an overlap of consideration across documents, a content saturation was reached.

**Table S2:** g characteristics of researchers involved in the content analysis

|  | JMS | LA |
| --- | --- | --- |
| Credentials | MD, BSc (Geography) | MSc (Epidemiology),<br>MA (Health Promotion),<br>BA (Social Work) |
| Occupation | Research scientists and PhD<br>Student Epidemiology and<br>public health at LMU Munich | Advisor epidemiology and health<br>monitoring at the Academy of Public<br>Health Services in Düsseldorf |
| Gender | Male | Female |
| Experience and training | Has received training on<br>qualitative content research at<br>the University of Tübingen<br>(Germany);<br>Multiple completed research<br>projects employing qualitative<br>content analysis. | Has received training on qualitative<br>content research at the University of<br>Applied Science Ravensburg-Weingarten (Germany); Participation<br>and implementation of various<br>research projects with qualitative<br>content analysis. |

#### 10.2.4 Adapting the preliminary coding frame

Next, one researcher (JMS) conducted a thematic analysis of the passages assigned to the newly created codes as well as those passages noted down for expanding on or providing nuance within existing codes. A draft of an adapted phase II framework was created by reflecting on whether there was a need to adapt the *a priori* framework to cover the content in the coded passages. This included whether: (a) *criteria* (reflecting level 1 codes) should be created, (b) new aspects (reflecting level 2 codes) should be added to the framework, (c) new or preexisting *criteria* or *aspects* should be merged or separated, or (e) moved to another position within the framework, and (f) wording of the *criteria* or *aspects* needs to be adapted.

This newly developed draft of the adapted framework was critically reviewed by a second researcher (LA). Afterwards, the results of steps (a-f) were critically discussed within the research team (JMS, LA) to solve conflicts and revise the adapted phase II framework accordingly.

Afterwards, two researchers jointly reviewed all passages noted down for later review (code TBD) and assessed the need to add *criteria* or *aspects*, as well as the need for revision, rewording or repositioning within the framework.

### Supplement 2: *a priori* framework adapted in phase I and coding frame

**Table S3:**
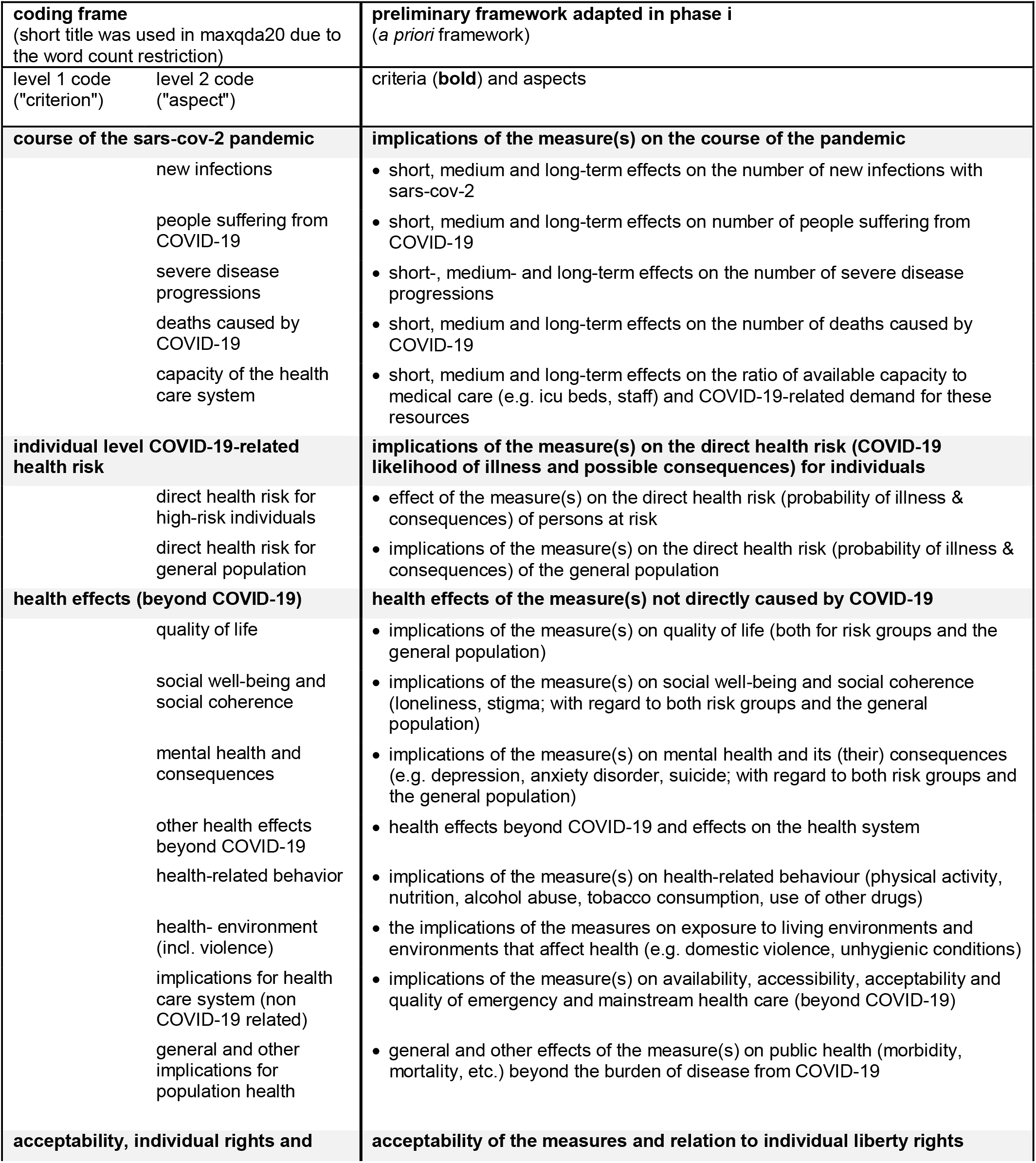
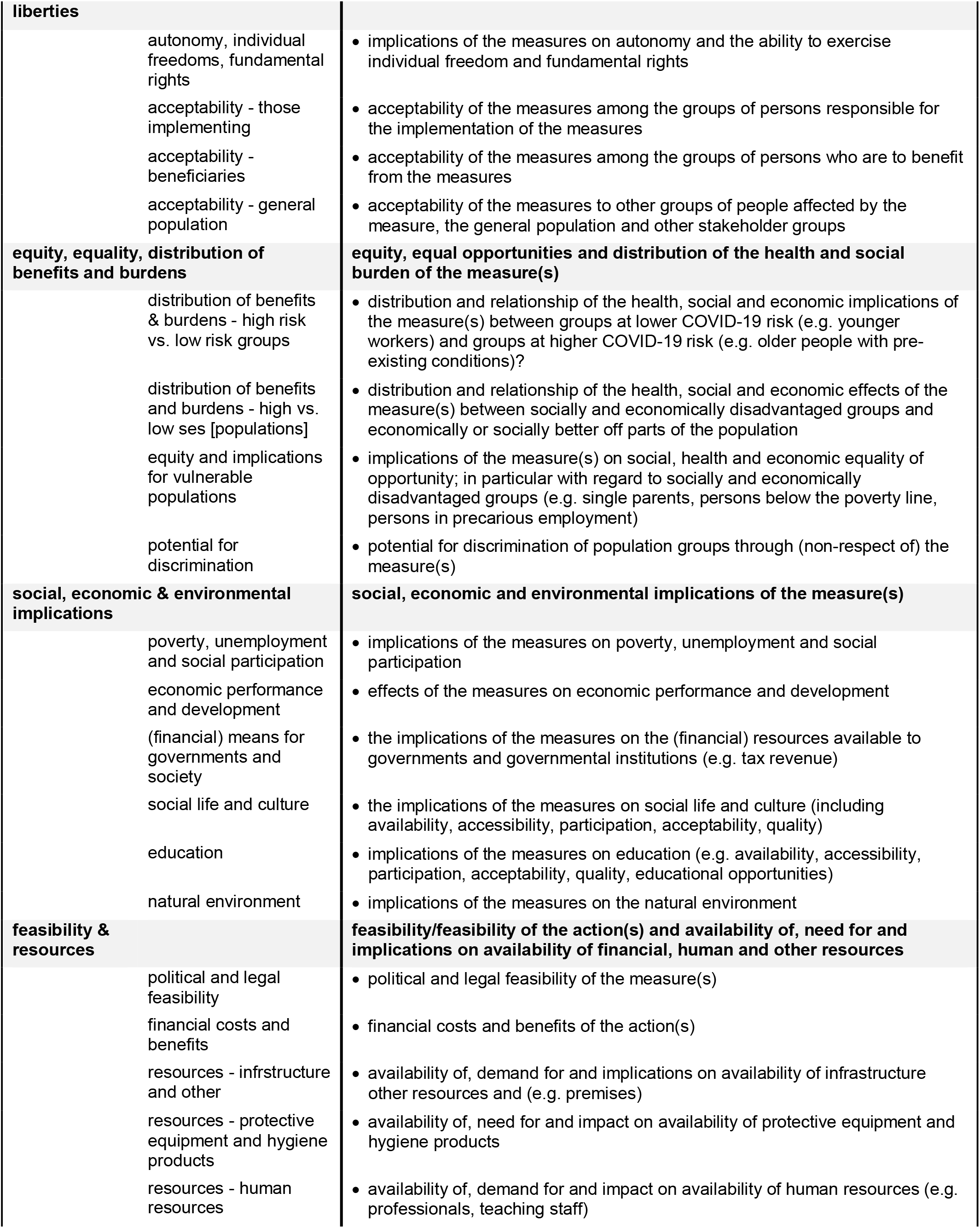
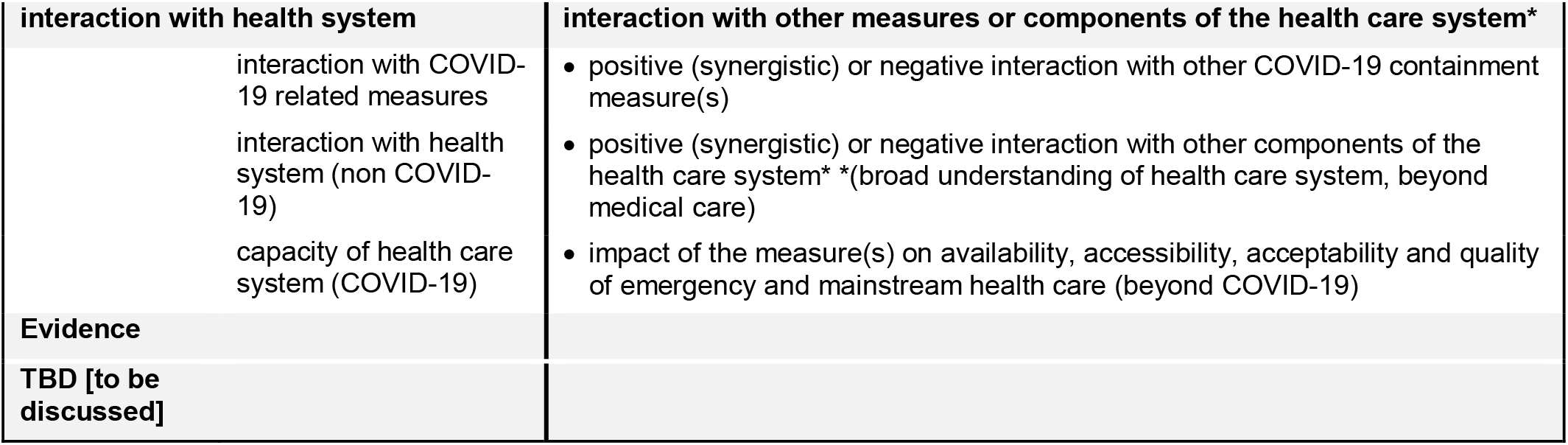
a priori framework adapted in phase I and coding frame The right column presents the preliminary adapted framework of the WHO-INTEGRATE framework, which was developed in phase I. It was used as the a priori framework for the “best fit” framework synthesis of phase II and translated into a coding frame (left two columns)

### Supplement 3: WICID Framework – comprehensive Version 1.0

**Table S4:**
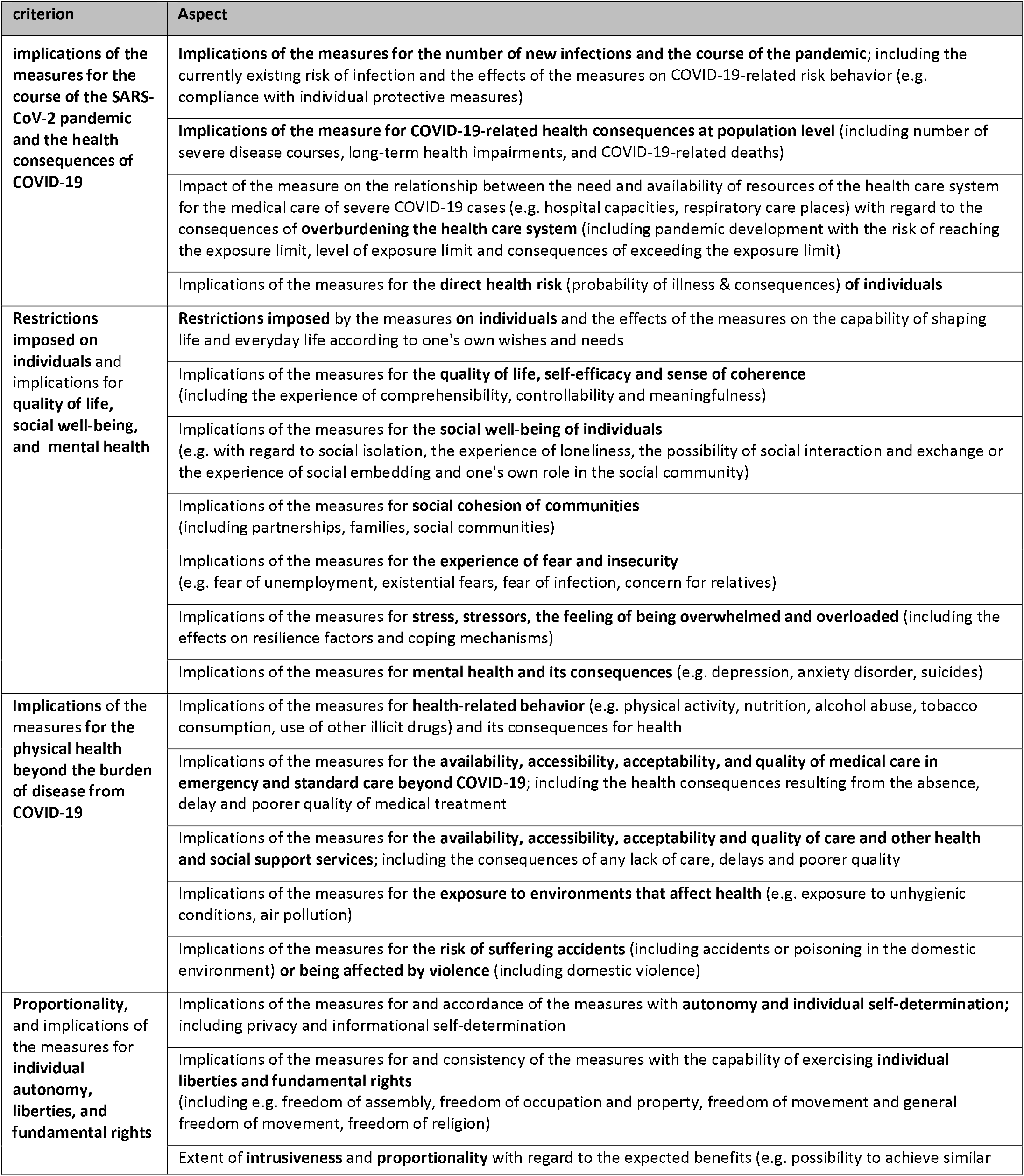
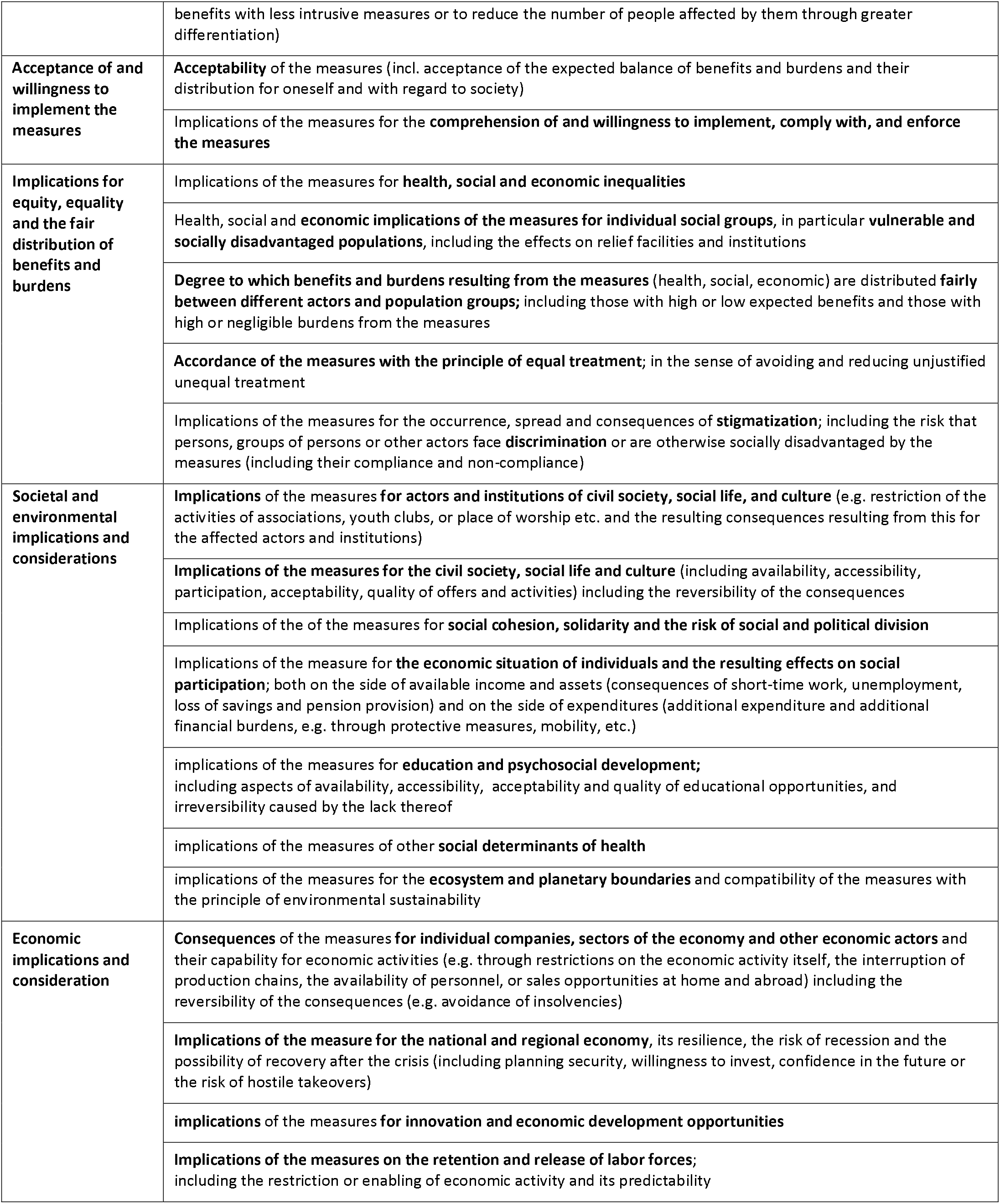
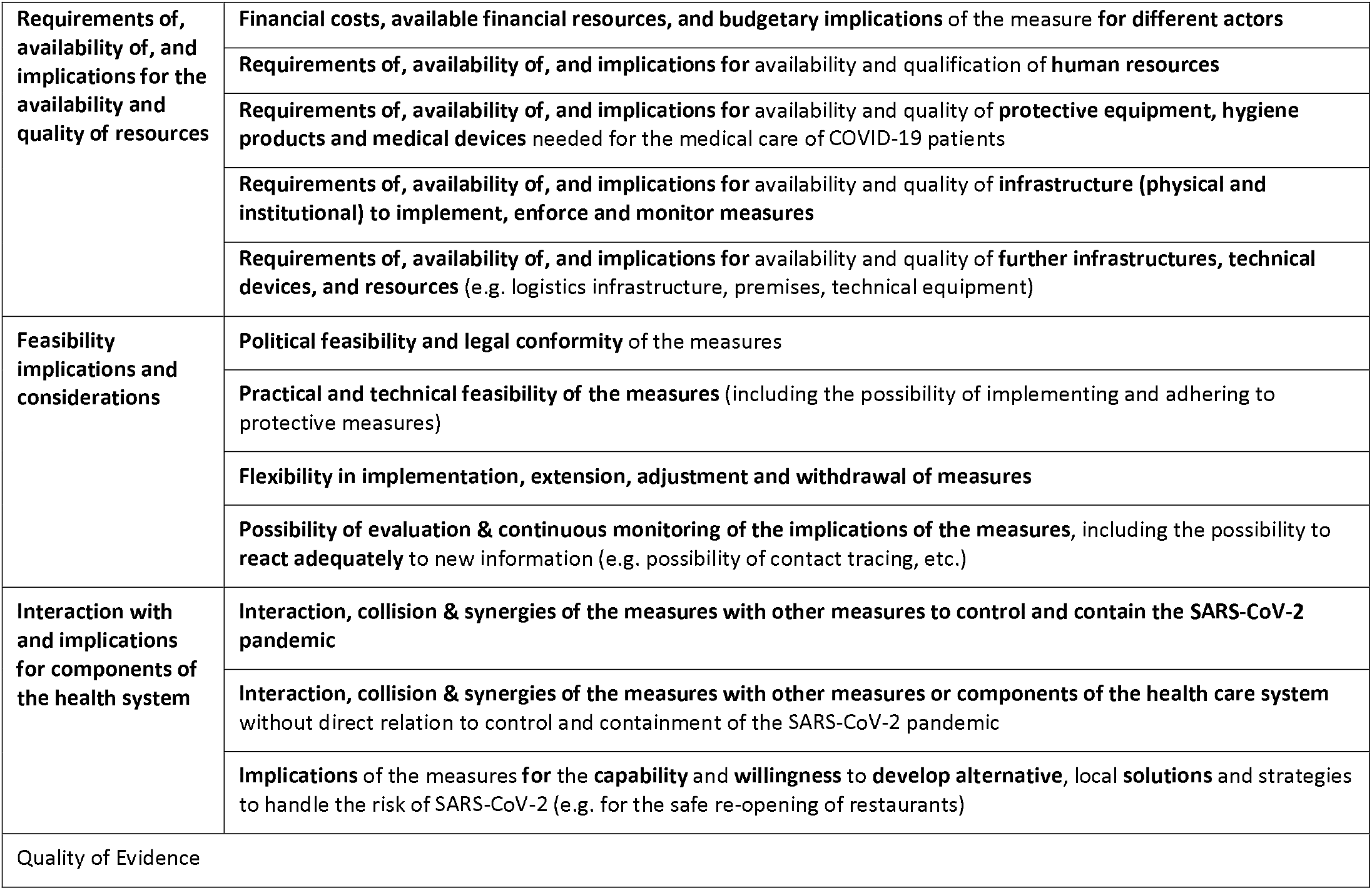
Criteria and aspects within (comprehensive form).

### 10.3 Supplement 4: Table with exemplary passages for the criteria and aspects of the WICID framework

**Table S5:**
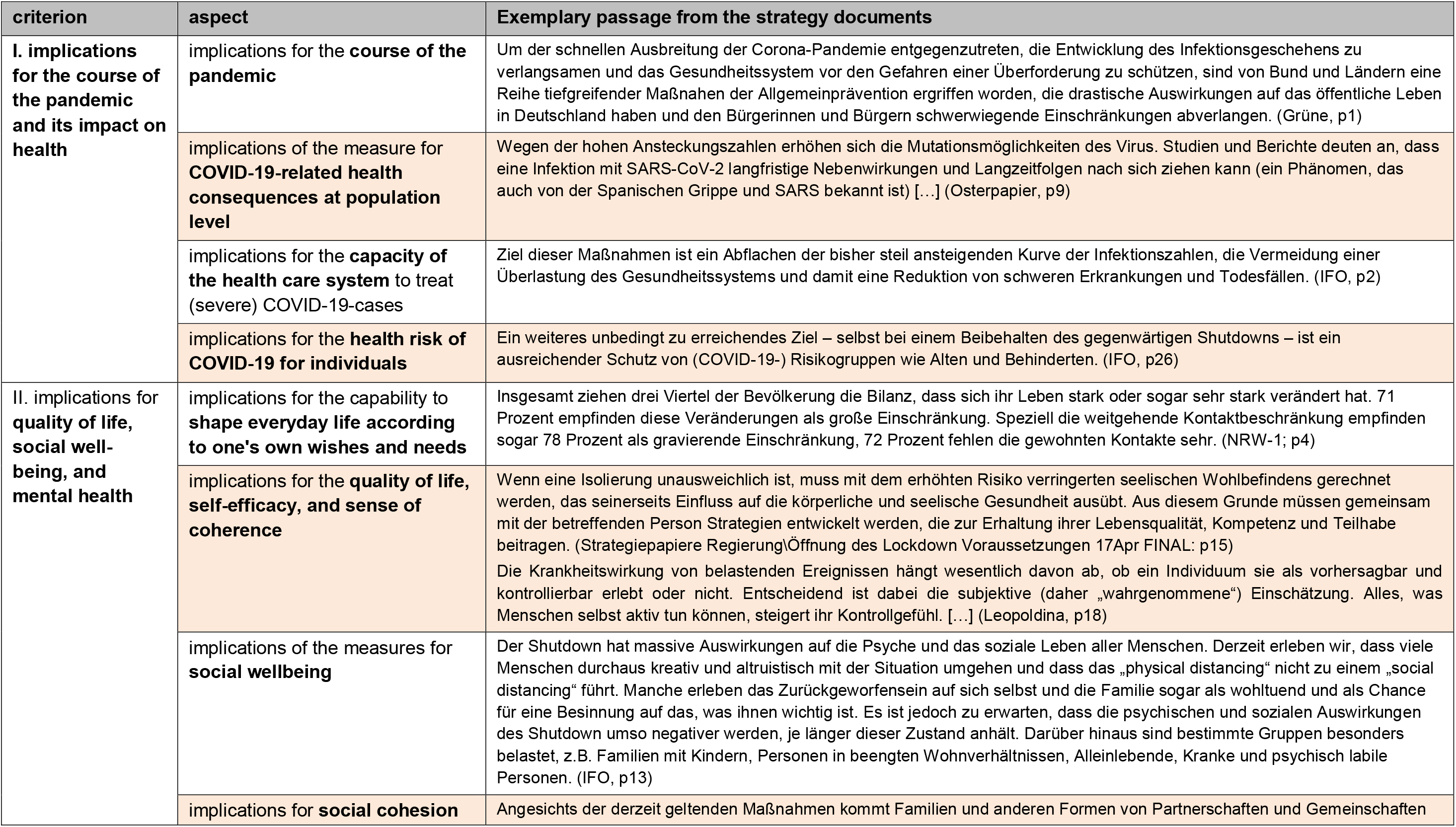
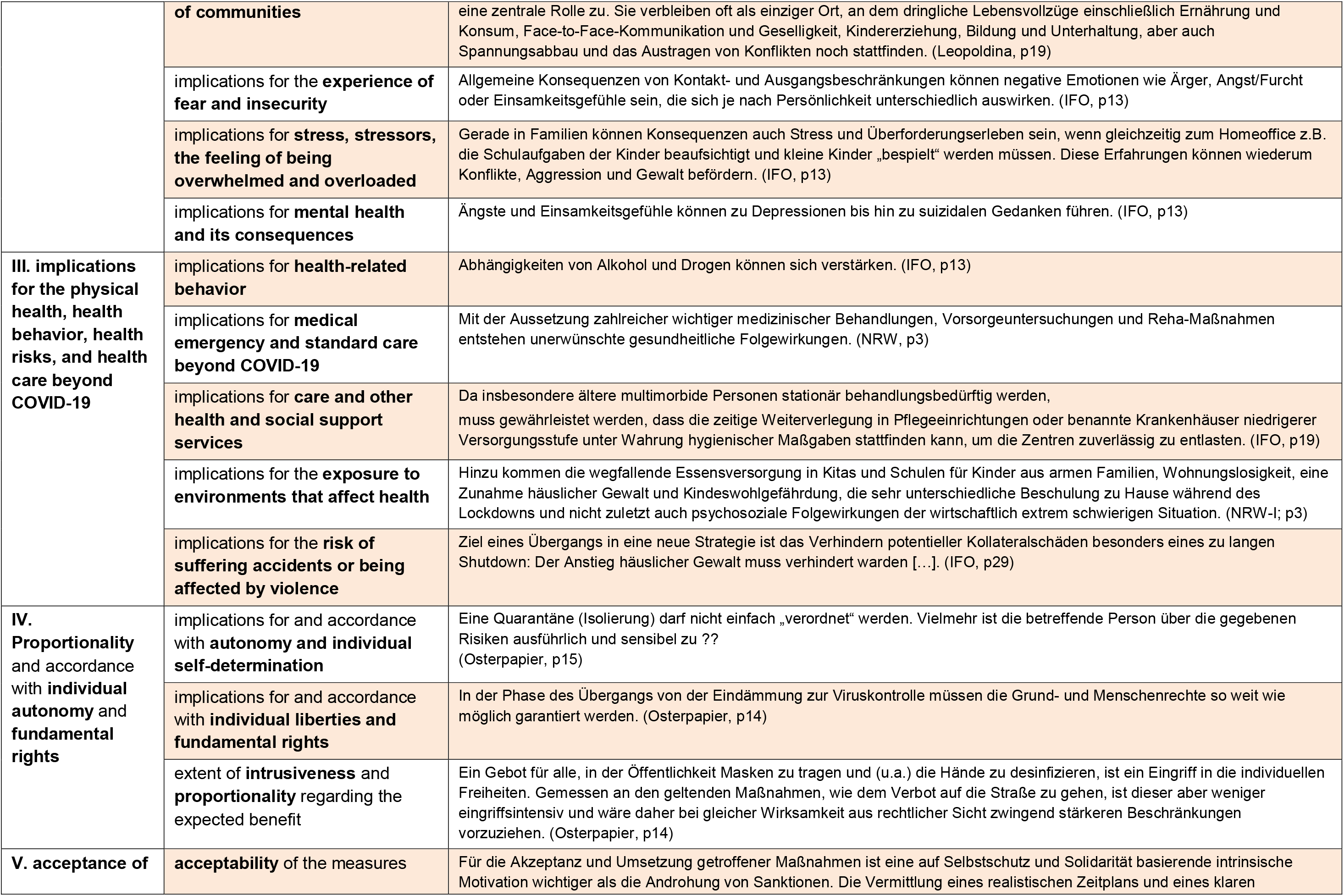
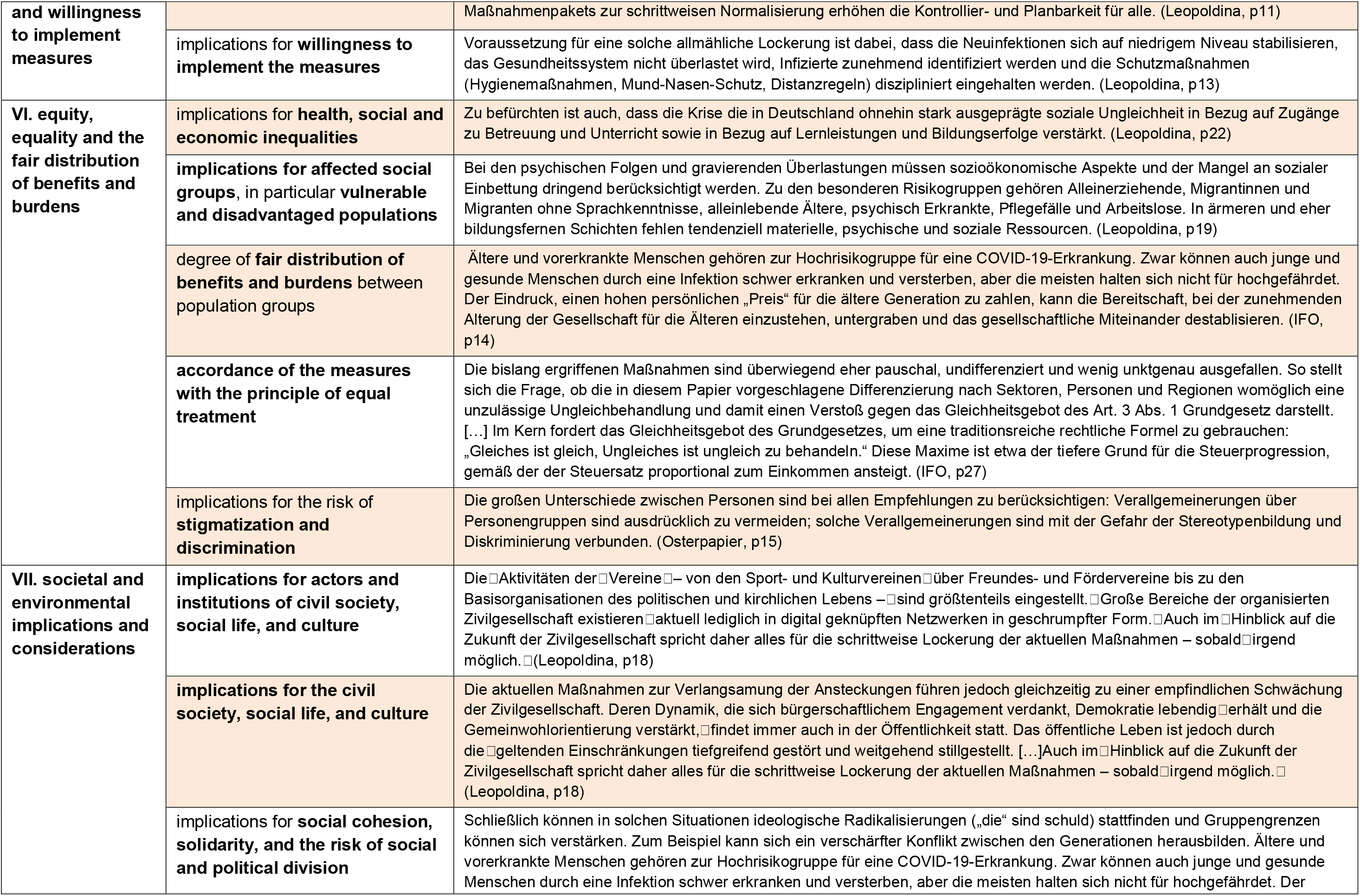
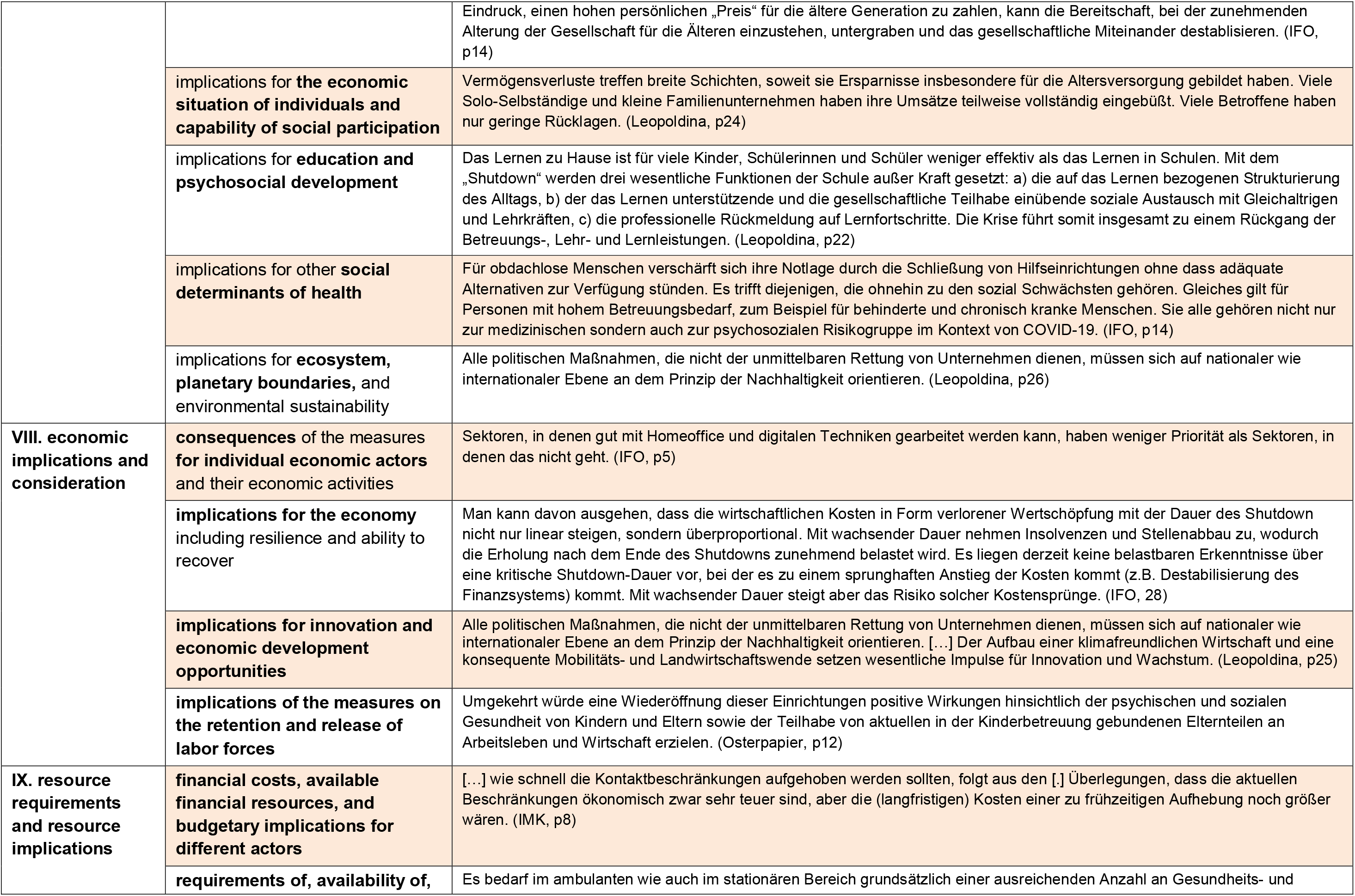
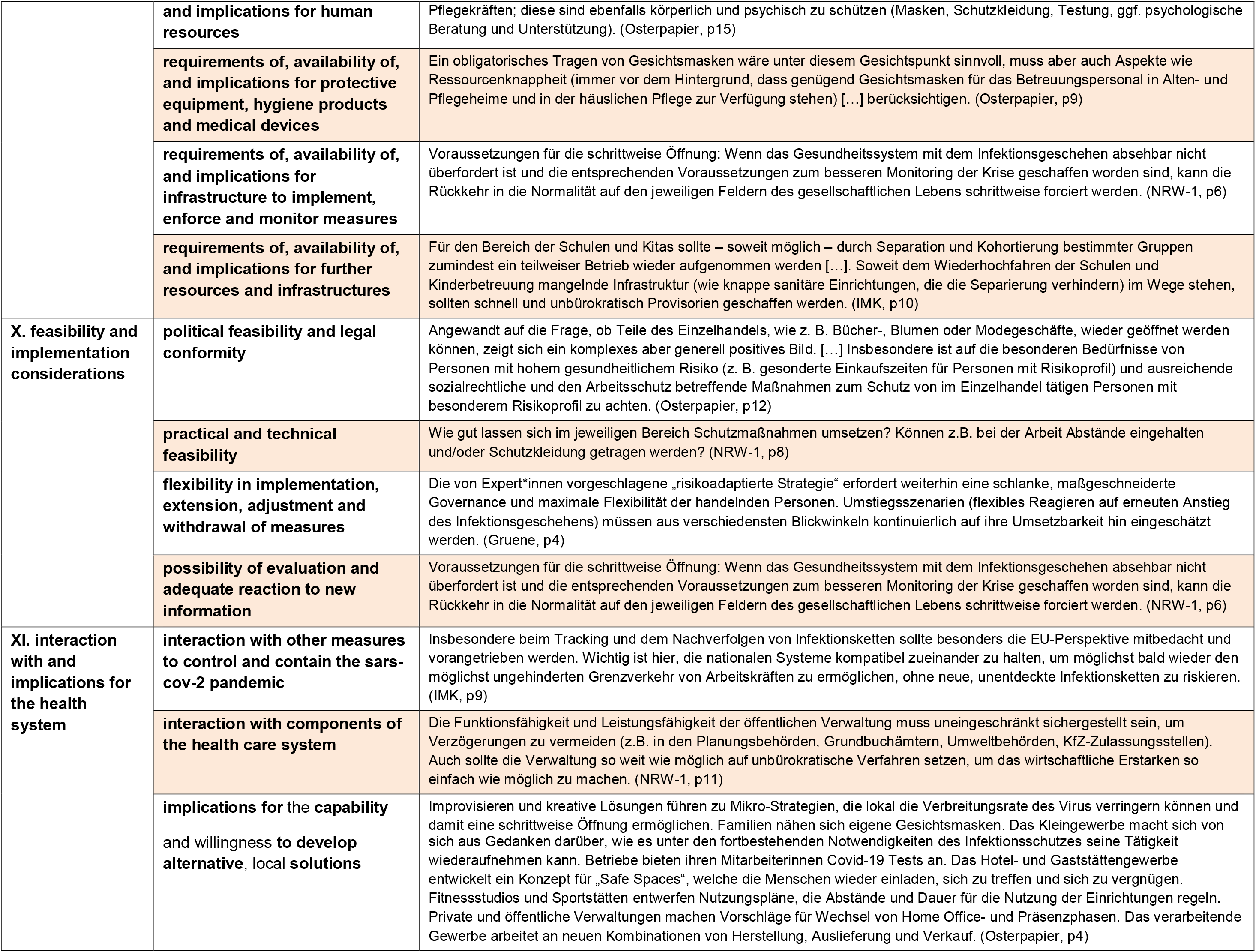
Criteria and aspects of the WICID framework with exemplary codes from the trategy documents

